# Molecular identification of SARS-CoV-2 variants of concern at urban wastewater treatment plants across South Africa

**DOI:** 10.1101/2022.12.15.22283506

**Authors:** Mukhlid Yousif, Said Rachida, Setshaba Taukobong, Nkosenhle Ndlovu, Chinwe Iwu-Jaja, Wayne Howard, Shelina Moonsamy, Nompilo Mhlambi, Sipho Gwala, Joshua I. Levy, Kristian G. Andersen, Cathrine Scheepers, Anne von Gottberg, Nicole Wolter, Arshad Ismail, Melinda Suchard, Kerrigan McCarthy, the SACCESS network

**Affiliations:** Centre for Vaccines and Immunology, National Institute for Communicable Diseases, a division of the National Health Laboratory Service, South Africa; Department of Virology, School of Pathology, Faculty of Health Sciences, University of the Witwatersrand, Johannesburg; Department of Immunology and Microbiology, The Scripps Research Institute, La Jolla, CA 92037, USA; SAMRC Antibody Immunity Research Unit, Faculty of Health Sciences, University of the Witwatersrand, Johannesburg, South Africa; Centre for Respiratory Diseases and Meningitis, National Institute for Communicable Diseases, a division of the National Health Laboratory Service, South Africa; School of Pathology, Faculty of Health Sciences, University of the Witwatersrand, Johannesburg, South Africa; Sequencing Core Facility, National Institute for Communicable Diseases, a division of the National Health Laboratory Service, South Africa; Department of Biochemistry and Microbiology, Faculty of Science, Engineering and Agriculture, University of Venda, Thohoyandou, South Africa; Department of Chemical Pathology, School of Pathology, University of the Witwatersrand, Johannesburg; School of Public Health, University of the Witwatersrand, Johannesburg

**Keywords:** environmental, wastewater, mutations, sequence, whole genome sequencing, SARS-CoV-2, COVID-19, surveillance, minor, spike, alpha, beta, delta, omicron, C1.2

## Abstract

The use of wastewater for SARS-CoV-2 surveillance is a useful complementary tool to clinical surveillance. The aims of this study were to characterize SARS-CoV-2 from wastewater samples, and to identify variants of concern present in samples collected from wastewater treatment plants in South African urban metros from April 2021 to January 2022. A total of 325 samples were collected from 15 wastewater treatment plants. Nucleic acids were extracted from concentrated samples, and subjected to amplicon-based whole genome sequencing. To identify variants of concerns and lineages, we used the Freyja tool (https://github.com/andersen-lab/Freyja), which assigns each sample with the prevalence of each variant present. We also used signature mutation analysis to identify variants in each wastewater treatment site. A heatmap was generated to identify patterns of emerging mutations in the spike gene using Excel conditional formatting. Using the Freyja tool, the Beta variant was detected and became predominate from April to June 2021 followed by the Delta variant and lastly the Omicron variant. Our heatmap approach was able to identify a pattern during the changes of predominate variant in wastewater with the emergence of mutations and the loss of others. In conclusion, sequencing of SARS-CoV-2 from wastewater largely corresponded with sequencing from clinical specimens. Our heatmap has the potential to detect new variants prior to emergence in clinical samples and this may be particularly useful during times of low disease incidence between waves, when few numbers of positive clinical samples are collected and submitted for testing. A limitation of wastewater sequencing is that it is not possible to identify new variants, as variants are classified based on known mutations in clinical strains.

## Background

As SARS-CoV-2 is shed into stool and urine, and is detectable in wastewater ^1^, quantification and sequencing of SARS-CoV-2 in wastewater has the potential to overcome inherent limitations in clinically-based epidemiological approaches. Over the pandemic, clinical surveillance has relied on testing and sequencing of samples from infected individuals. However, when clinical testing forms the basis for surveillance, population health seeking behaviour, test accessibility and testing practices of attending clinicians limit the generalisability of data. In particular, only symptomatic patients approach the health system for testing and testing practices vary by location and over time ^2^, leading to an incomplete representation of local virus spread and diversity. Testing of wastewater for SARS-CoV-2 levels overcomes these limitations by allowing population levels of SARS-CoV-2 to be monitored over time and by location, adding key information to our understanding of SARS-CoV-2 transmission dynamics. The value of wastewater monitoring of SARS-CoV-2 is attested to by the fact that over 70 countries now provide monitoring and public reporting of geographical and temporal trends in wastewater levels ^3 4^

Wastewater genomic surveillance offers an opportunity to monitor circulating variants present in the community. Whole genome sequencing ^5^, and other methods such as real-time PCR ^6^ enable detection and characterisation of SARS-CoV-2 variants, and have now been applied to wastewater samples. Recent work has shown the potential for recovery of complete virus genomes from wastewater^7^, shown comparable results of wastewater and clinical surveillance, and identified novel mutations and lineages before appearance in clinical samples ^8 2^. To date, wastewater sequencing of SARS-CoV-2 has not been widely applied in low or middle income countries.

South Africa is a middle-income country with a population of over 55 million persons, most of whom live in urban centres located in five of the country’s nine provinces. South Africa has over 1,000 wastewater treatment plants ^9^ and the majority of South Africans (84%) have access to piped sanitation (flush toilets connected to a public sewerage system or a septic tank) ^10^. Wastewater testing for SARS-CoV-2 was first described in South Africa in June 2020 ^11^. The South African Collaborative COVID-19 Environmental Surveillance System (SACCESS) arose to monitor trends in SARS-CoV-2 levels in wastewater ^12^.

Most laboratory testing is provided to the public through an extensive network of laboratories including the National Health Laboratory Service (NHLS) that covers over 80% of the population. South Africa identified its first case of COVID-19 on the 5^th^ of March 2020, ^13^ and four waves of COVID-19 occurred within the first 24 months of virus introduction into the country. Following the initial SARS-CoV-2 wave, the Beta variant ^14^ was discovered and was predominant from November 2020 to February 2021 (second wave). The third wave (May to September 2021) was characterized by the dominance of the Delta variant ^15,16^ and the fourth wave (November 2021 to January 2022) by the Omicron BA.1 variant ^17^. The National Institute for Communicable Diseases (NICD), a division of the NHLS, provides SARS-CoV-2 epidemiological surveillance data through collation of SARS-CoV-2 PCR results from public and private laboratories. The Network for Genomics Surveillance in South Africa (NGS-SA) monitors the epidemiology of SARS-CoV-2 variants in PCR-confirmed cases in South Africa and reports weekly on findings ^18^. The NGS-SA provided the first global reports of the emergence of Beta and Omicron variants of concern (VOC) ^14,17^.

Here, we show that wastewater can be used to effectively characterize SARS-CoV-2 virus dynamics in the population and identify variants of concern present using samples from sentinel wastewater treatment plants in urban metros collected from April, 2021 to the end of the fourth wave in January, 2022, demonstrating the utility of wastewater genomic surveillance to complement clinical surveillance efforts in a middle income setting. We identify the potential strengths and limitations of genomic wastewater surveillance for SARS-CoV-2 in the South African context.

## Methods

### Wastewater sites

A total of 325 samples were collected from 15 wastewater treatment plants (WWTP) in metropolitan areas also undergoing quantitative analysis of SARS-CoV-2 by the NICD ^19^. Sites were situated in Gauteng, Eastern Cape, Western Cape, Free State, and KwaZulu-Natal provinces. Supplementary table

S1 shows population sizes draining to each WWTP, and number of samples collected and sequenced. The samples were collected between April 2021 to January 2022 during the third and fourth waves of SARS-CoV-2 infections in South Africa.

### Sample collection, RNA extraction, amplification and sequencing

One liter of grab sewage samples were collected and transported to NICD at 4°C. Viruses were concentrated from the sample by ultrafiltration ^20^, and RNA was extracted using the QIAamp Viral RNA kit (Qiagen, GmbH, Germany). SARS-CoV-2 was detected by RT-PCR using the Allplex™ 2019-nCoV Assay from Seegene (Seoul, Korea). RNA was re-extracted from SARS-CoV-2 positive concentrates and subjected to amplicon-based whole genome sequencing using the Sinai protocol with some modifications as described ^21 22^. Paired-end libraries were prepared using Illumina COVIDSeq Kit as previously described ^23^ followed by sequencing (2x 150 pb) on NextSeq 1000/2000 platofrm (Illumina Inc, USA).

### Sequence analysis

#### Quality control checks

FastQ files were trimmed, filtered based on sequence quality, assembled and mapped to the reference genome (NC_045512.2) according to published criteria ^24^ using Exatype web-based bioinformatics tool (https://sars-cov-2.exatype.com/). Quality control indicators such as number of reads, number of mapped reads, and sequence coverage were recorded. Samples that passed the internal quality control were processed for mutation analysis using ARTIC protocol (https://artic.network/ncov-2019/ncov2019-bioinformatics-sop.html) in Galaxy (https://usegalaxy.eu/) ^25^. Again, reads were trimmed and filtered, assembled and mapped. At least 10 reads were required at each nucleotide position for downstream analysis. Amino acid mutations present at 5% of reads or less were removed from the analysis. Table 1 illustrates an example of amino acid variation analysis output.

**Table 1.** Illustration of amino acid variations in samples, produced by the Galaxy pipeline.

| A | B | C | D | E | F | G |
| --- | --- | --- | --- | --- | --- | --- |
| Sampl<br>e | QC filter | Numb<br>er<br>of<br>reads | Mutation effect | GENE | Mutatio<br>n | Mutatio<br>n<br>frequen<br>cy |
| 21-0128 | PASS | 4073 | NON_SYNONYMOUS_COD<br>ING | ORF1a<br>b | E41K | 92 |
| 21-0128 | PASS | 12106 | NON_SYNONYMOUS_COD<br>ING | S | L1203F | 96 |
| 21-0128 | min_af_0.1Xmin_<br>dp_5Xmin_dp_alt<br>_10 | 12234 | FRAME_SHIFT | S | V1228 | 1 |
| 21-0128 | PASS | 13569 | NON_SYNONYMOUS_COD<br>ING | ORF3a | T190I | 88 |
| 21-0128 | PASS | 1536 | NON_SYNONYMOUS_COD<br>ING | E | P71L | 97 |
| 21-0128 | min_af_0.1Xmin_<br>dp_5Xmin_dp_alt<br>_10 | 1339 | NON_SYNONYMOUS_COD<br>ING | M | I8S | 1 |

**A** shows sample ID. **B** is the quality indicator. Filter value 'PASS' are samples with a minimum of 10 reads and read frequency greater than 10. **C** is the number of reads produced for each sample. **D** is the effect of the mutation detected in the gene. **E** is the name of the gene where mutation occurred. **F** is the mutation detected. **G** is the proportion of the mutant cells in a sample or population.

### Sequence analysis using amino acid mutations

#### Amino acids variation analysis

As SARS-CoV-2 RNA in wastewater is fragmented, and fragments originate from multiple individuals (generally infected with genetically distinct viruses), the generation of consensus sequences from wastewater samples is not meaningful. Rather, we inferred the presence of variants by using amino acid mutations uniquely associated with each VOC, as follows: Using the amino acid variation data file generated by the Galaxy pipeline above, we used STATA software (v 17.1) (https://www.stata.com/) to collate spike-gene mutations in a matrix such that the columns represented the amino acid positions of the spike protein and each row recorded mutations identified from a single wastewater sample. We included all mutations, and recorded the proportion of reads where that mutation was detected (the ‘read frequency’) as a percentage of total reads. For each VOC or Variant of Interest (VOI), we identified signature single amino acid mutations by comparing the new variant/ lineage with the Wuhan reference sequence in a public database ^26^ (Table 2). Using this list of unique mutations for each VOC and VOI in the spike protein region (Table 2) we interrogated our matrix for the presence or absence of known signature mutations in each sample using STATA software. As new variants/lineages were detected and identified in clinical specimens, we added signature mutations to the STATA code, allowing us to identify the presence of new variants both retrospectively and prospectively.

**Table 2:** List of signature mutations which was used to identify VOC and VOI present in wastewater samples (Gangavarapu *et al*., 2022)

| Omicron | Alpha | Beta | Delta | C.1.2 | Gamma | Lambda | Mu |
| --- | --- | --- | --- | --- | --- | --- | --- |
| G339D | A570D | D80A | T19R | P9L | T20N | G75V | Y144S |
| S371L | S982A |  | R145H | P25L | P26S | T76I | Y145N |
| S375F | D1118H |  | E156del | C136F | T1027I | D253N |  |
| Q493R |  |  | R158G | Y449H |  | L452Q |  |
| G496S |  |  | A222V |  |  | F490S |  |
| Y505H |  |  |  |  |  |  |  |
| T547K |  |  |  |  |  |  |  |
| N856K |  |  |  |  |  |  |  |
| Q954H |  |  |  |  |  |  |  |
| N969K |  |  |  |  |  |  |  |
| L981F |  |  |  |  |  |  |  |

#### Heatmap and dot blot

Using the amino acid variations data output file from Galaxy and the generated excel file that contains all amino acid variations and their respective read frequency, a heatmap was generated to identify patterns of emerging mutations in the spike gene using Excel conditional formatting. To identify the events and times at which uncommon mutations were identified in our samples, an in-house R script (R v.4.2.0) was used to generate a mutational dot plot.

#### Analysis using the Freyja tool

To capture the dynamics of virus evolution and spread, we used Freyja ^27^, a tool to estimate the relative abundance of virus lineages present in wastewater. Freyja uses a “barcode” library of lineage-defining mutations to uniquely define all known SARS-CoV-2 lineages and solves for lineage abundance using a depth-weighted, least absolute deviation regression approach. Freyja is free to use and available at https://github.com/andersen-lab/Freyja.

## Results

### Quality control

A total of 325 wastewater samples from sites listed in Table S1 were amplified and sequenced. The median number of sequence reads was 1.72×10^6^ (inter quartile range [IQR] 2.53×10^5^-2.67×10^6^) with a range of 6.5×10^4^ to 8.16×10^6^reads (Figure S1). A total of 229 (70.5%) samples had > 1 million reads, and 237 (72.9%) samples had >50% reads mapped to the reference sequence (range 0-89%). Regarding sequence coverage in 10x depth, 183 (56.3%) samples had >50% sequence coverage of the whole genome (Figure S2a), and 177 (54.5%) samples had >50% sequence coverage in the spike region (Figure S2b).

### Detection of SARS-CoV-2 variants from wastewater samples using signature mutation analysis

Signature mutations (Table 2) were identified in 170 samples (52.3%), 79 samples from Gauteng, 32 from KwaZulu-Natal, 32 Free State, 12 from Western Cape, and 15 from the Eastern Cape provinces respectively. Figure 1 illustrates the signature mutations for VOCs that were identified in samples from each wastewater treatment plant. In most of wastewater treatment plants a transition change in VOCs was observed in all wastewater treatment plants in the country, from Beta to Delta to Omicron during post second wave, third wave of fourth wave of infections. C.1.2 was also detected during Delta wave.

**Figure 1.**
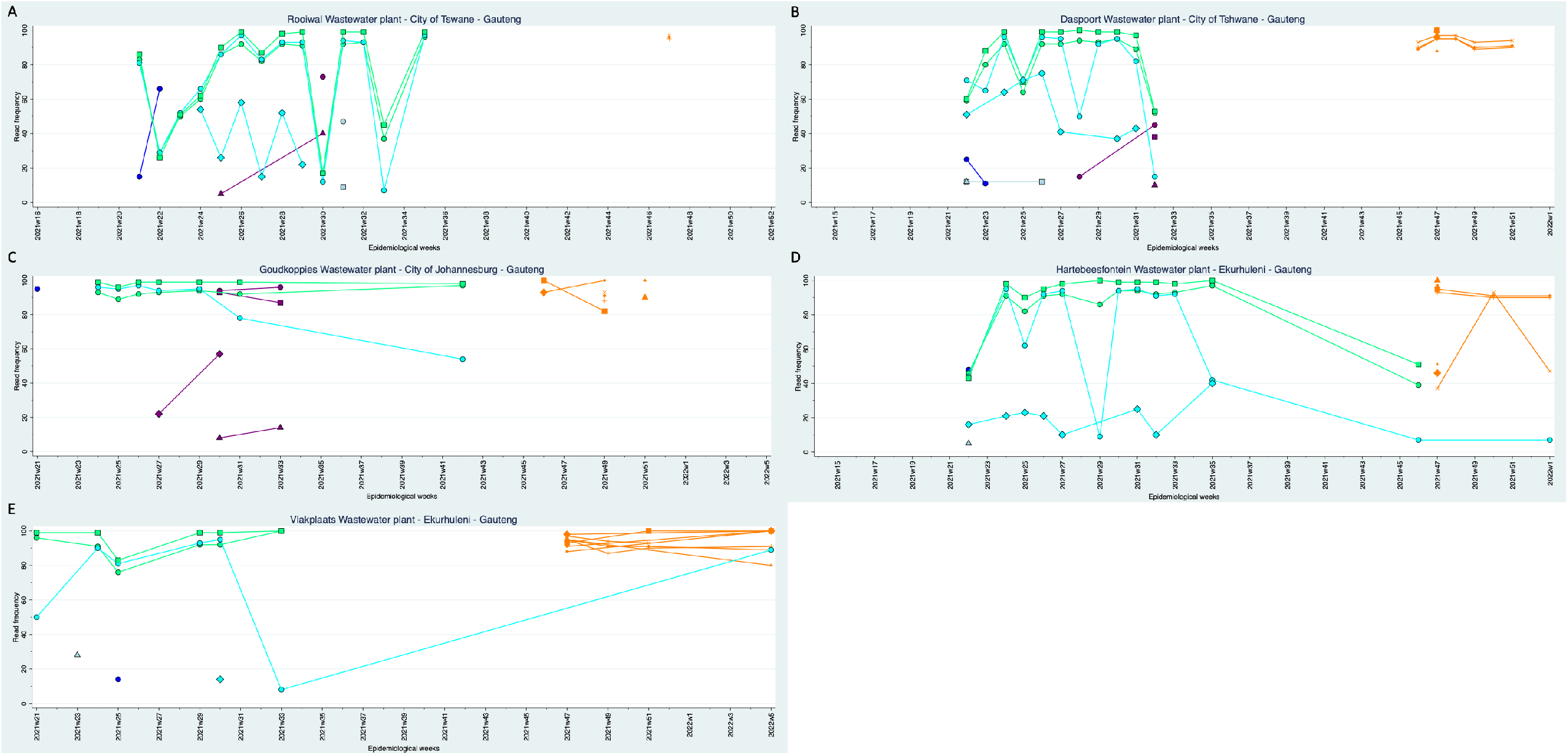
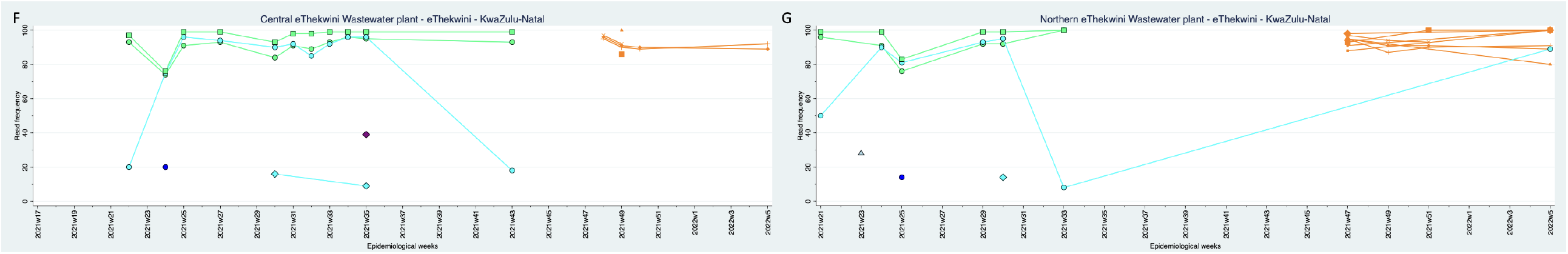
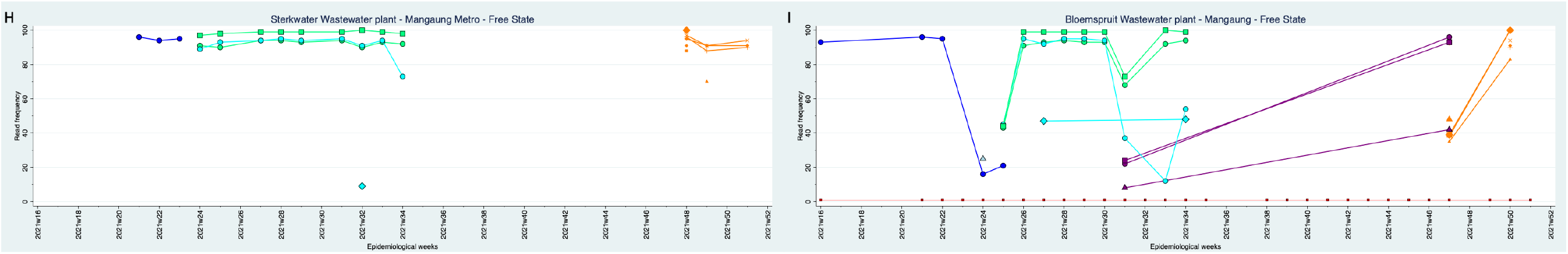
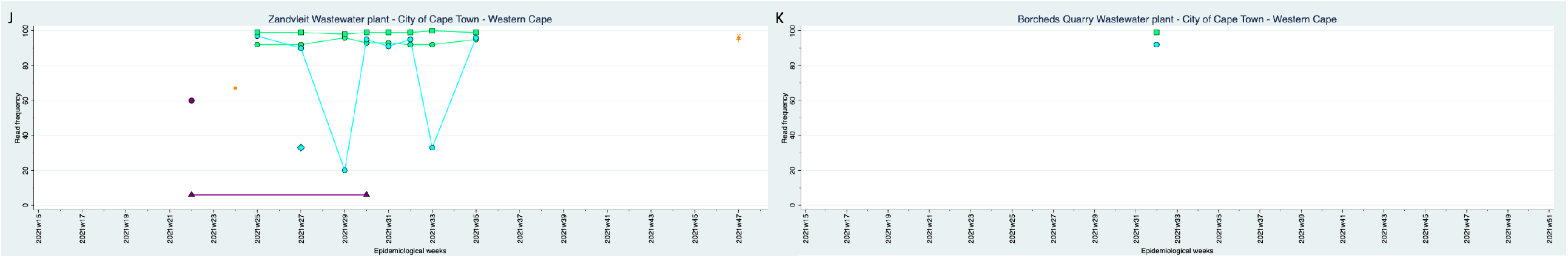
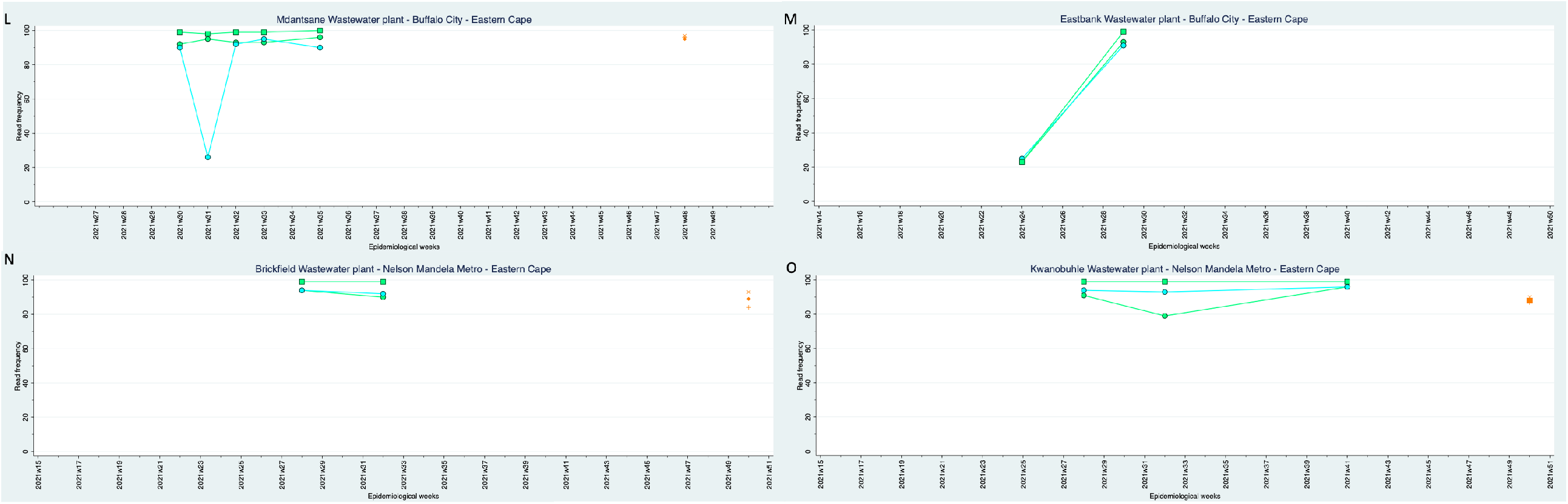
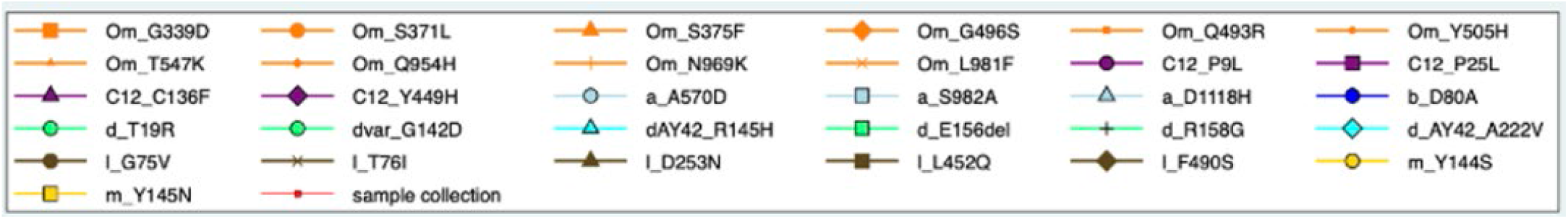
Signature mutation analysis of Variants of concern (VOC) and Variants of interest (VOI). The Y axis shows the read frequencies of each mutations, and X axis shows the epidemiological week when the samples were collected. The red line with dots shows the time-point analysed. Different shapes and colours of signature mutations were shown in the key. A-E: Gauteng province sites. E-G: KwaZulu-Natal province. H-I: Free State province. J-K: Western Cape province. L-O: Eastern Cape province. P: key for the different shapes and colours for each mutation and variants.

### Detection of SARS-CoV-2 variants from wastewater samples using Freyja tool

Out of the 325 samples sequenced, 168 (51.7%) samples had a sequence coverage of >50% at 10x depth, and were successfully assigned a SARS-CoV-2 variant using the Freyja tool (Figure 2). In these samples, the Beta variant was detected and became predominate from April to June 2021. Delta variant emerge in May 2021 and predominates in June until October 2021. Omicron variant was detected in November 2021 and immediately dominate until January 2022. Alpha variant was seen in small proportion in June and July 2021 and lineage A and variant Kappa was detected in June 2021 in a very small proportion (Figure 2a). The proportions of lineages were shown in Figure 2b, a total of 68 lineages were detected of which 53 lineages were also reported in clinical cases and 15 lineages were not reported in clinical cases.

**Figure 2.**
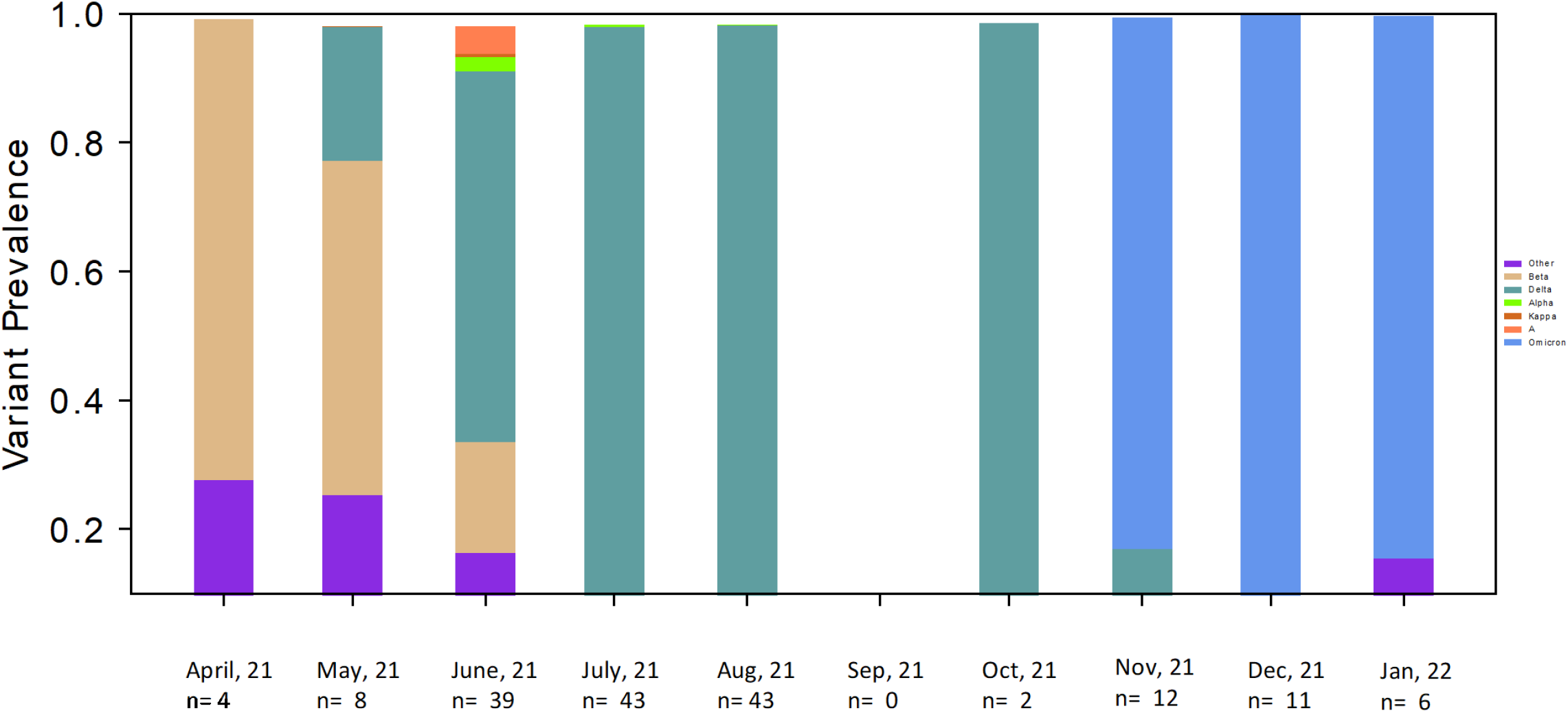
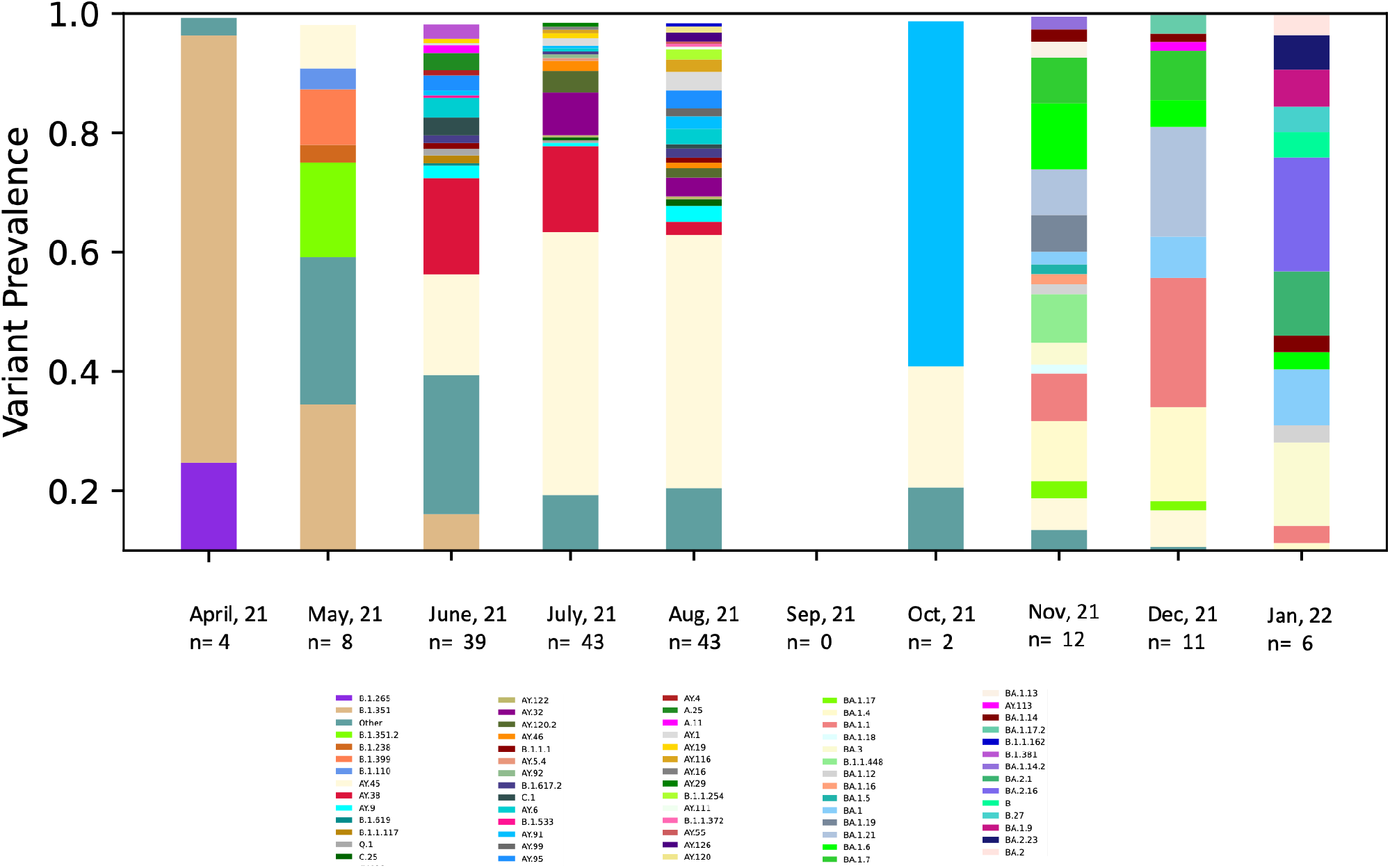
The proportion of variants and lineages by month from wastewater samples, from April 2021 to January 2022 using the Freyja tool. The X-axis shows the month and the number of samples sequenced. The Y axis shows the proportion of each variant present in the specific month. Only samples with sequence coverage of >50 were included. “Other” represents unconfirmed lineages found in the wastewater samples. **A**. SARS-CoV-2 variants. **B**. SARS-CoV-2 lineages

### Characterization of amino acid mutations in the spike region

A total of 411 amino acid mutations were observed in the spike protein amongst all sequenced samples. Alignment by amino acid position in a heatmap (Figure 3) demonstrated a characteristic pattern of mutations in each epidemiological wave of COVID-19. The transition from Delta variant to Omicron was characterized by a loss of mutations in the N-terminal domain (NTD) region (E156del, F157del, and R158G), and new mutations in the receptor binding (RBD) domain (G339D, S371L, 373, N440K, S477N, E484A, Q493R, G496S, Q498R), and fusion peptide (FP) region (N764K, D796Y), and the heptad repeat 1 (HR1) region (Q954H, N969K, L9811F). Between the third and fourth wave of infection low sequence coverage of spike was observed, likely due to low caseload, and few mutations were detected. Of the 411 substitutions/ deletions detected, 78 were present at >1% prevalence. When we compared those mutations to known published mutations at GISAID (https://gisaid.org/), 58 were commonly reported (Table S2), and 10 were uncommonly reported (Figure 4).

**Figure 3.**
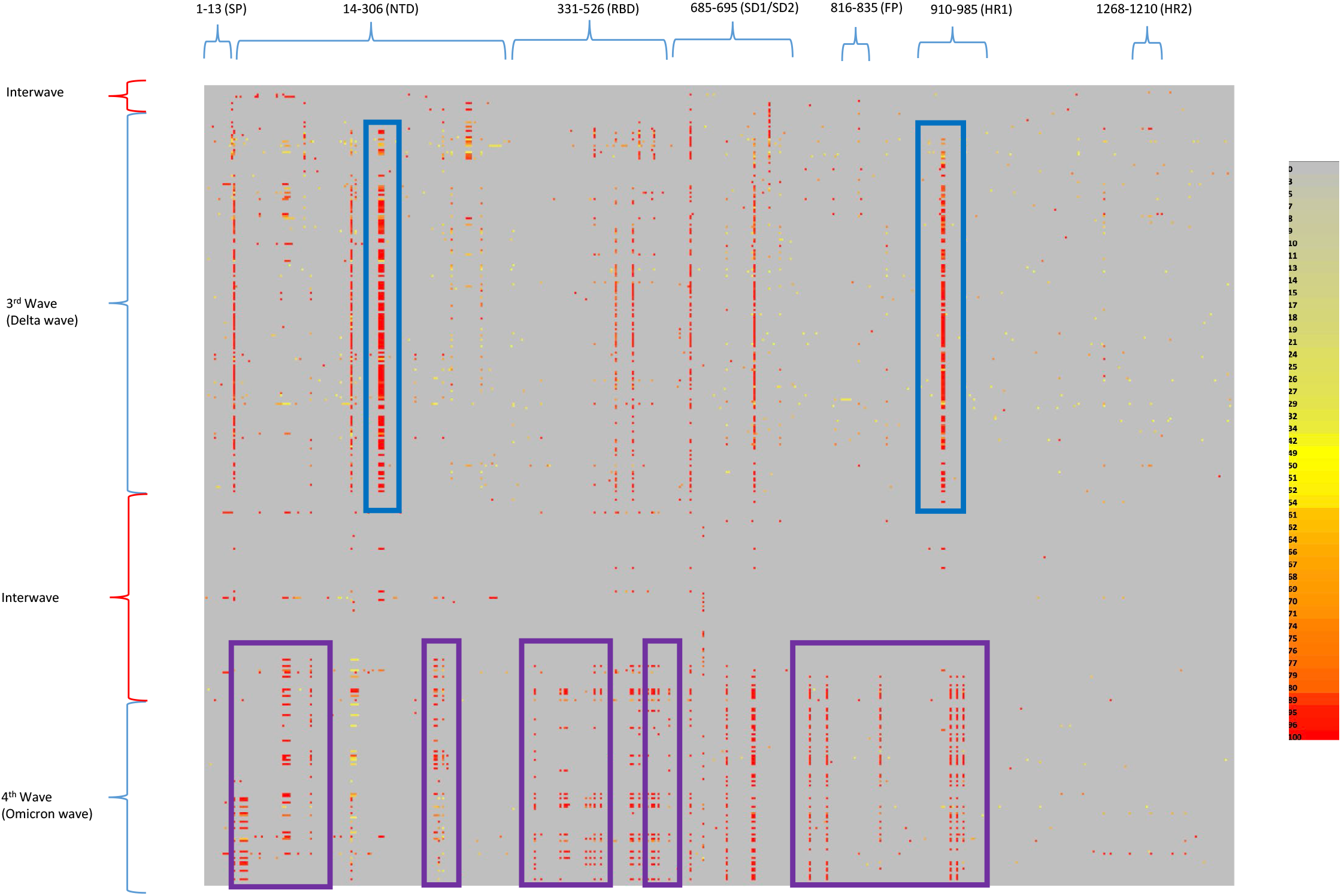
Heatmap of amino acid mutations distributed across the SARS-CoV2 spike protein in comparison with the Wuhan reference strain, arranged vertically in chronological order. Each row represents a sample, organized by the date of sample collection. Each column represents an amino acid position of the spike protein. Regions with no mutations or low occurrences are represented in grey and light yellow (0-34%). Regions with mutations that have a 50% read frequency are represented in bright yellow. Regions with mutations with a read frequency between 60-80% are represented in orange and very high occurring mutations (89-100%) are represented in red, as per the key.

**Figure 4.**
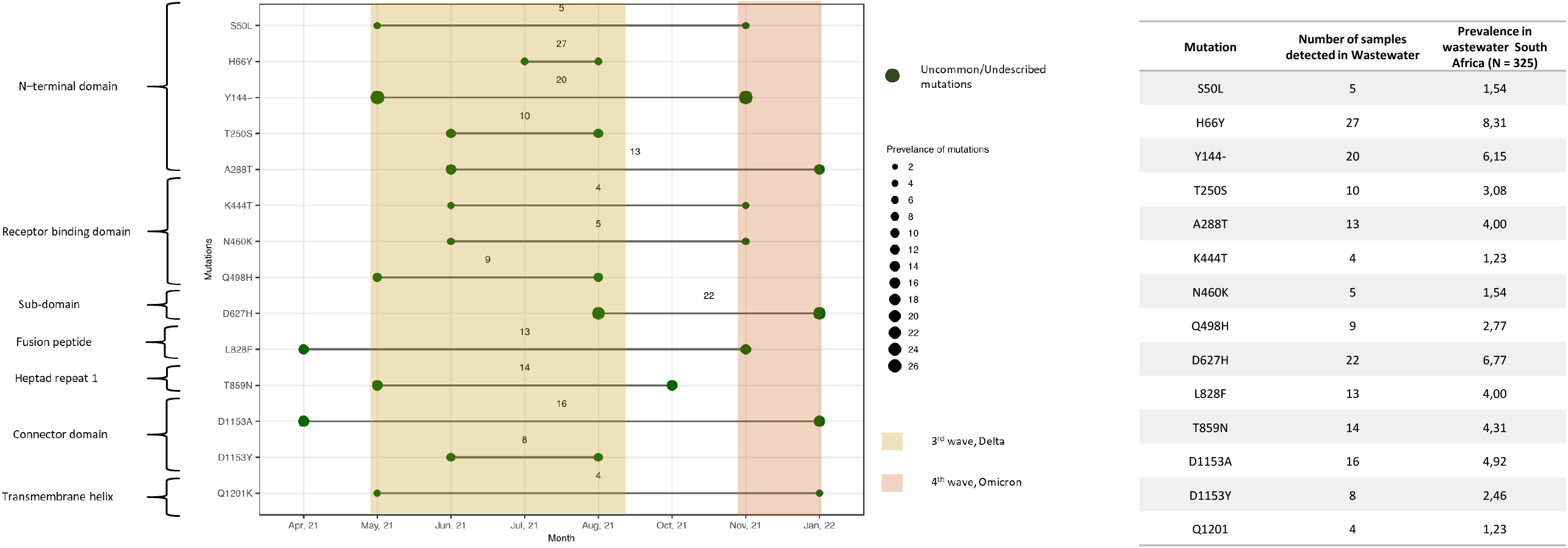
Dot blot showing the uncommon mutations of SARS-CoV-2 detected in wastewater during the period April, 2021 – January, 2022 and their prevalence. The x-axis represents the uncommon mutations and the y-axis represents the times at which the mutations were first and last observed (as represented by continuous line between the dates). The size of the dot of each mutation describes the number of times the mutation was observed (prevalence) in the collected samples.

## Discussion

In this study, our sequencing methodology and bioinformatics pipeline facilitated detection and genomic characterization of SARS-CoV-2 lineages present in South African wastewater treatment plants in five provinces during the period April 2021 to January 2022. Our results showed the presence of all VOCs also detected amongst clinical samples (Beta, Delta, and Omicron). We detected amino acid mutations that were well-described and available publically in the GISAID database, and also amino acid mutations that were uncommon or rarely reported in clinical samples. We were also able to show the emergence of new mutations and loss of other mutations during the time of wave changes from Beta to Delta and from Delta to Omicron. Collectively, these findings illustrate how sequence analysis of SARS-CoV-2 in wastewater complements epidemiological findings based on clinical sequencing.

Wastewater is a complex matrix containing highly fragmented virus genomic material. Our methodology generated a high number of reads with good quality and depth when compared to other studies that have sequenced SARS-CoV-2 from wastewater^28^. In addition, the highest proportion of mapped reads was as 89% which was in the range of what have been achieved in other studies ^2^. The highest sequence coverage in a depth of 10x reads in this study was 99% for the whole genome and spike protein.

Our approach to interpretation of sequence data generated from wastewater samples is unlike analysis of sequences derived from clinical specimens. Because wastewater contains a mix of RNA fragments from viral particles originating from many infected individuals, the generation of consensus sequences is not meaningful. Our approach allows for identification of previously described variants in wastewater samples and also for detection of new patterns of mutations suggesting previously undescribed variants. Similarly, this approach has also been used Crits-Chritoph and colleague ^8^.

Regarding detection of previously described variants in our wastewater samples, both approaches we used (mutational analysis and the Freyja tool) successfully identified lineages in wastewater that corresponded to lineages identified in clinical specimens ^18^. The Beta variant (first described in clinical specimens in South Africa in December 2020 ^14^) was consistently observed from the start of wastewater surveillance until the end of the third wave in epidemiological week 19 (May 2021). Similarly, the Delta variant was first seen in clinical specimens during epidemiological week 21 of 2021 ^16^, and was first detected in wastewater samples the same week. Lineage C.1.2, described first in South Africa ^29^ was successfully detected in sequences from wastewater samples during week 22 to 45 in 2021 whilst clinical detections of this lineage appeared from week 16 to 46. In epidemiological week 46 in 2021, the Omicron variant was identified in clinical samples whilst sequences from wastewater samples also identified mutations specific to Omicron in the same week. The Freyja tool ^30^ complemented our mutational profile analysis, identifying the proportions of each variant/lineage in wastewater at specific time points. Results from Freyja comparable to the prevalence of VOCs of SARS-CoV-2 reported from clinical specimens and additionally indicate the presence of lineages in our wastewater samples that were absent amongst sequences from clinical cases. This most likely arises through the sampling bias inherent in clinical surveillance, in which only symptomatic patients are tested and of whom only a fraction was sequenced.

Regarding detection of previously undescribed variants, our use of a spike-protein heatmap illustrated how each variant had a distinct mutational profile of RNA sequences of the spike gene, and that this changed in each wave. Through observation of the spike protein heatmap, samples with changing profiles may be identified before the new variant is sequenced from clinical isolates. This is clearly evident in the transition from the Delta to the Omicron variant, where a constellation of mutations in the NTD fell away and RBD, FP, and HR1 regions had new mutations (Figure 3). The shifting mutational profile correlated with the increased transmissibility of the Omicron variant that led to the fourth wave of infection in South Arfrica^17^.

Our mutational analysis identified multiple instances of rare mutations in the population (Figure 4). These mutations were found at a prevalence of >1.0% in wastewater samples, but were detected at <0.001% in clinical cases based on the data from GISAID. Although they were uncommon, some mutations were reported previously. Mutation S50L was found to be associated with reduced protein stability ^31^. Mutation Q498H has reportedly caused increased binding affinity of RBD to ACE2 ^32^. The presence of uncommon mutations could be explained as “cryptic lineages” as described by <u>Smyth</u> and colleagues ^33^. In their paper they attributed the cryptic lineages to either un-sampled cases or spillover of SARS-CoV-2 from an unidentified animal reservoir. Other wastewater studies have demonstrated cryptic variants, for example Yaniv and colleagues described a cryptic Delta variant in wastewater samples ^34^. Unusual substitutions /deletions are useful tools that support epidemiological tracking of variants and may support hypothesis generation regarding the origins of SARS-CoV-2.

Sequencing of SARS-Cov-2 in wastewater currently has a number of limitations. Refining methodological approaches is essential in order to prevent inhibition from substances within the wastewater matrix. Where the incidence of SARS-CoV-2 is very low, virus concentration in wastewater may proceed below the level of detection, which makes it difficult to amplify and sequence the viral genome. Further, emergence of a new variants, such as Omicron sub-variants BA.1 and BA.2, can lead to poor primer binding and lower coverage rates, particularly in the spike protein because of S gene dropout ^35^. Both of these scenarios renders sequencing of SARS-CoV-2 from wastewater challenging. In addition, bioinformatics methods for wastewater, including mutational analysis and the Freyja tool, are currently limited by their reliance on lineage assignment based on prior clinical sequencing and publicly available sequences.

## Conclusion

Sequencing of SARS-CoV-2 from wastewater largely corresponded with sequencing from clinical specimens. The prevalence of VOCs and lineages in clinical specimens was shown to be detectable in wastewater during the same times, which enabled us to provide comprehensive details on VOCs and lineages in the population. Despite inherent limitations of SARS-CoV-2 sampling in wastewater, we have generated a database spanning three SARS-CoV-2 waves, that document variant and lineage changes with time and geographical location and which correspond to clinically identified variants. We have illustrated how sequences not found in clinical specimens may be identified in populations through wastewater. Our heatmap has the potential to detect new variants prior to emergence in clinical samples and this may be particularly useful during times of low disease incidence between waves, when few numbers of positive clinical samples are collected and submitted for testing.

## Data Availability

All data produced in the present work are contained in the manuscript

## Declaration

### Ethics approval and consent to participant

The study did not involve any human participants. An application for ethics waiver was made to the Human Research Ethics Committee of the University of the Witwatersrand and was approved (number R14/49).

### Consent for publication

Not applicable

## Availability of data and materials

### Supplementary figures

**Figure S1** Stacked graph illustrating the total number of sequence reads from wastewater samples (N = 325) between April, 2021 – January, 2022. Mapped reads are represented in blue and unmapped reads are represented in orange. A horizontal red line inserted at the mark one million reads. Samples had sequence reads of more than one million were to the left of the red vertical line and samples had less than one million reads to the right of the red vertical line. The lower quartile value (2.53×10^5^), median (1.72×10^6^) and third quartile (2.6×10^6^) values are represented in blue horizontal broken lines.

**Figure S2**. A graph showing the percentage coverage of SARS-CoV-2 in 10x depth of sequence reads. Y-axis is the percentage of coverage, and x-axis is the total number of samples. **A**. sequence coverage for whole genome. **B**. sequence coverage for spike protein.

### Supplementary tables

**Table S1, table S2**,

## Authors contributions

MY: co-conceptualized study, co-performed analysis, wrote, edit, and reviewed manuscript. SR: co-conceptualized study, edit, and reviewed manuscript. ST: co-performed analysis, edit, and reviewed manuscript. NN: co-performed analysis, edit, and reviewed manuscript. CI: edit, and reviewed manuscript. WH: edit, and reviewed manuscript, SM: edit, and reviewed manuscript, SG: edit, and reviewed manuscript, JIL: co-performed analysis, edit, and reviewed manuscript, KGA: edit, and reviewed manuscript, CS: edit, and reviewed manuscript, AvG: edit, and reviewed manuscript, NW: edit, and reviewed manuscript AI: edit, and reviewed manuscript, MS: co-conceptualized, edit, and reviewed manuscript. KM: co-conceptualized, edit, and reviewed manuscript.

## Acknowledgements

The authors would like to thank the local government and wastewater treatment staff for sample collection and transport. We also thank the staff of the NICD Centre for Vaccines and Immunology and the Centre for Respiratory Disease and Meningitis. special thanks to: Josie Everatt, Boitshoko Mahlangu, Anele Mnguni, Noxolo Ntuli, Gerald Motsatsi for their assistance in setting up and troubleshooting PCR testing, and ongoing supportive collaboration. We thank the team at Hyrax Biosciences for the use of their tool exatype. We would like to acknowledge the contribution from the SACCESS network. We also acknowledge the funding from the NICD, Bill and Melinda Gates Foundation, Water Research Commission of South Africa, and Africa CDC.

