## Supplementary figures and images for "Molecular identification of SARS-CoV-2 variants of concern at urban wastewater treatment plants across South Africa"

### Figure S1

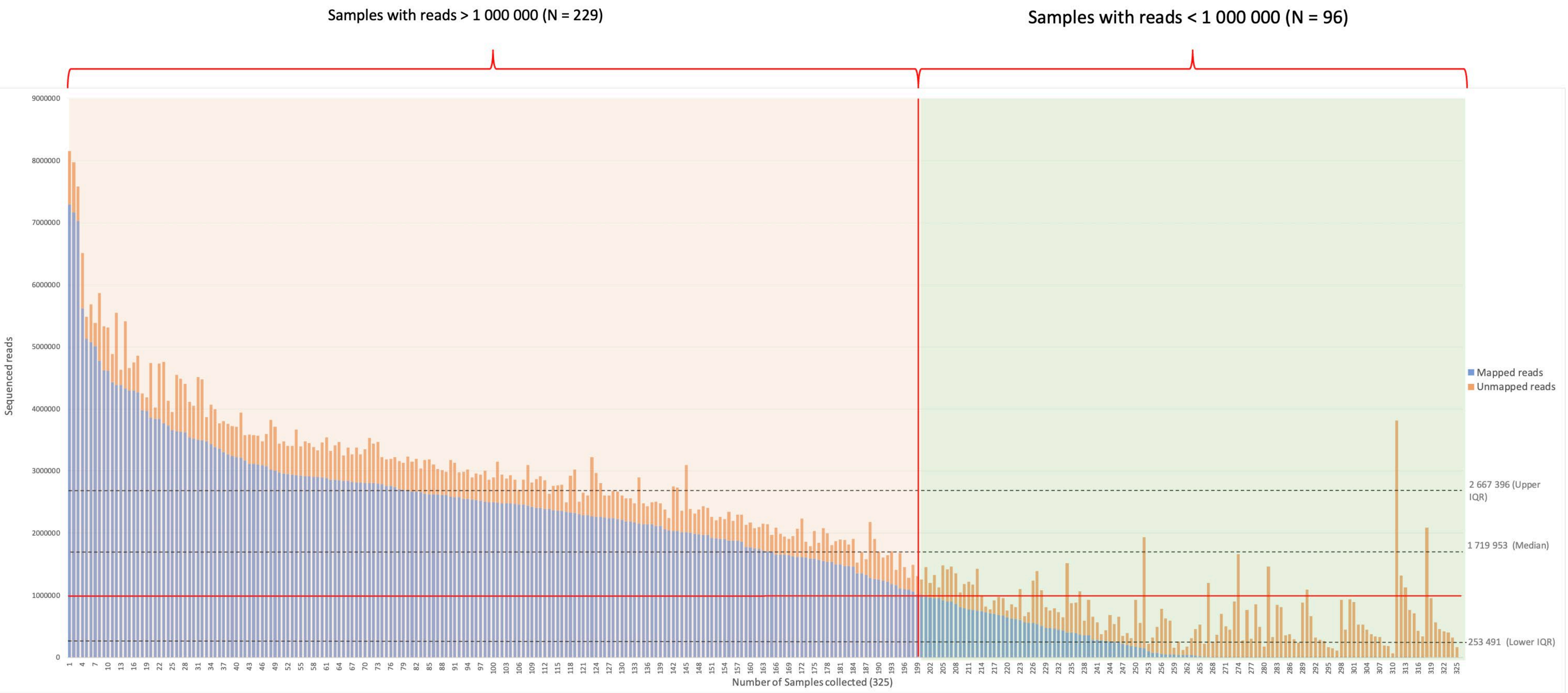

### Figure S2A

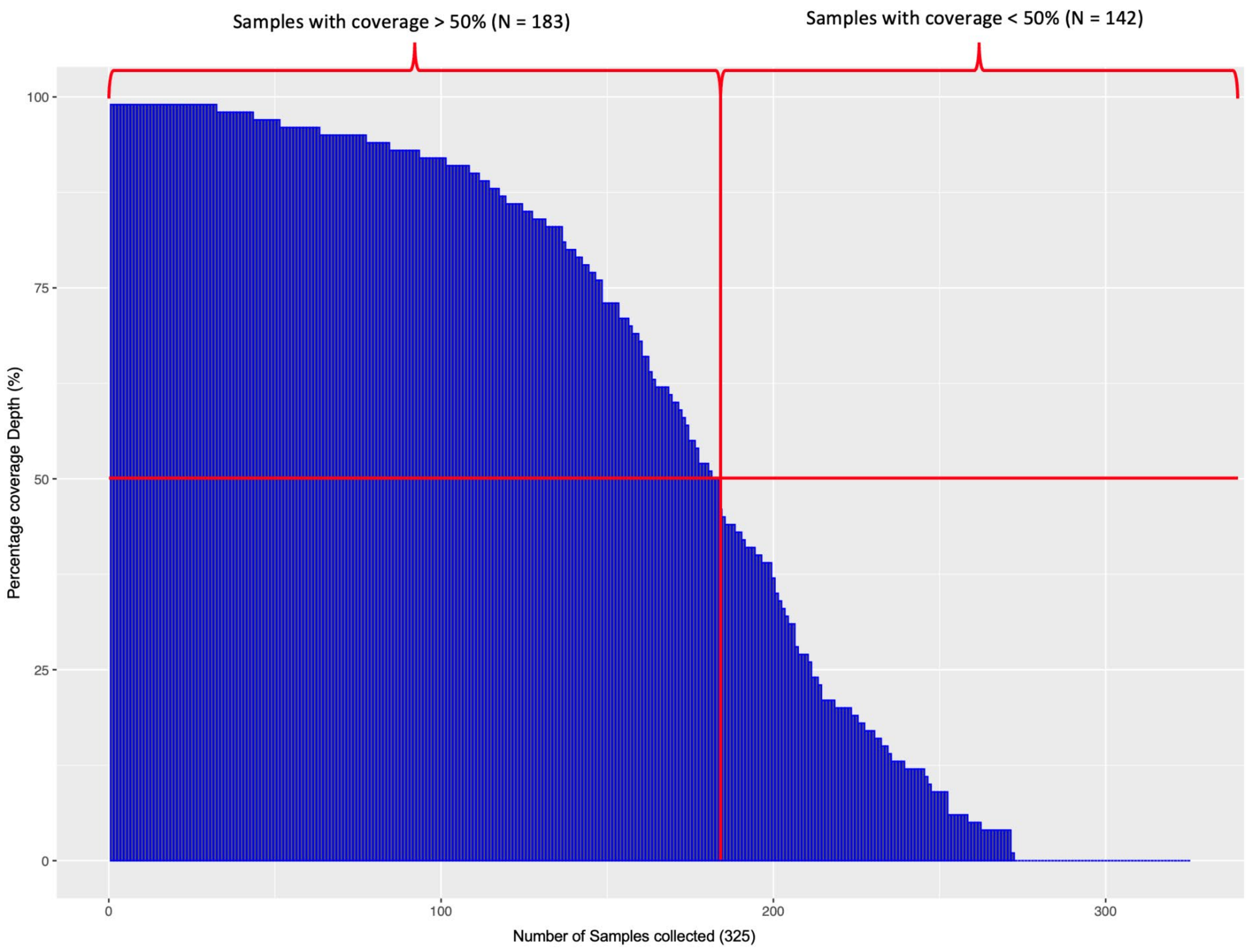

### Figure S2B

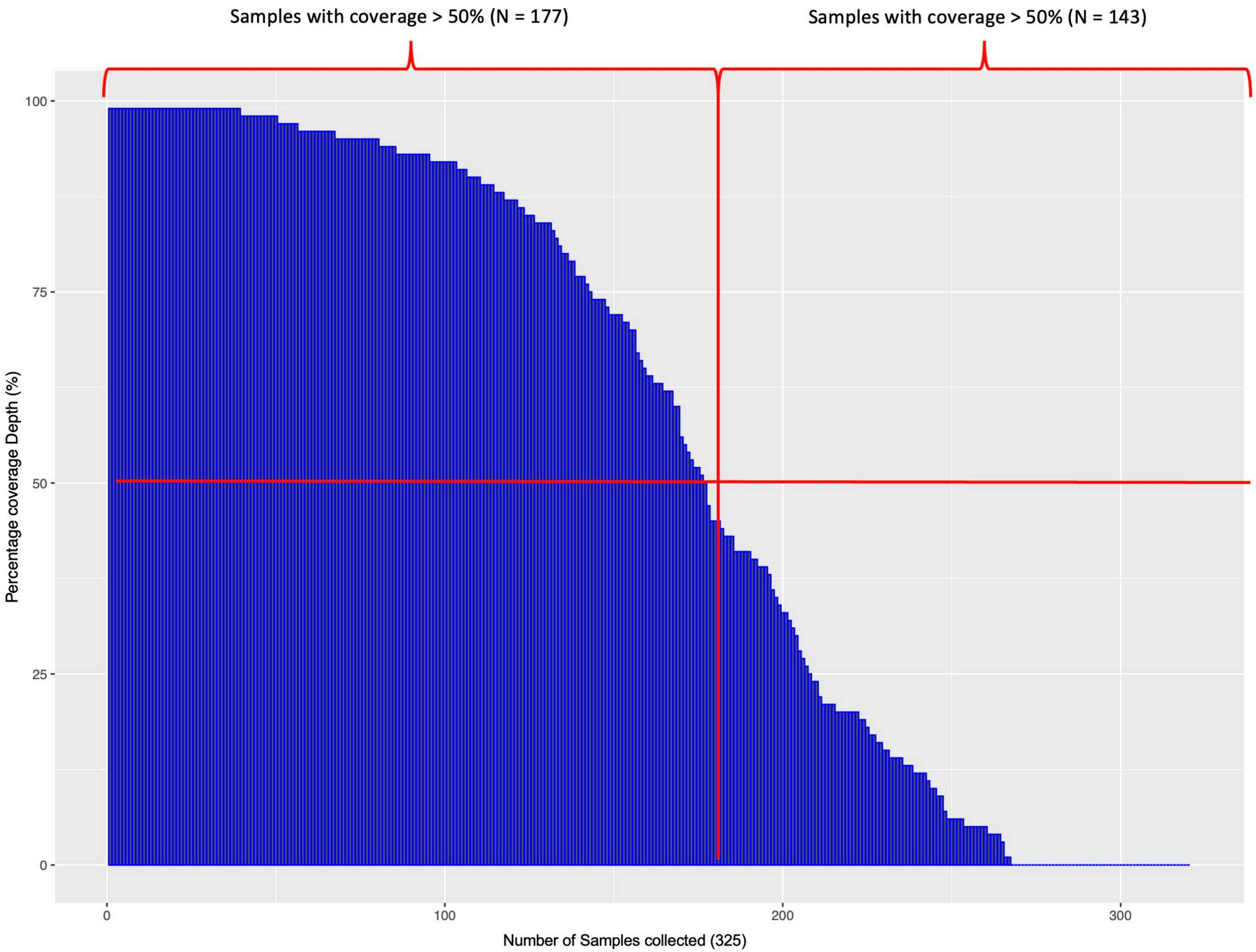
